# Critical success factors for routine immunization performance: A case study of Zambia 2000 to 2018

**DOI:** 10.1101/2021.11.30.21267060

**Authors:** Katie Rodriguez, Kyra A. Hester, Chama Chanda, Roopa Darwar, Bonheur Dounebaine, Anna S. Ellis, Pinar Keskinocak, Abimbola Leslie, Mwangala Manyando, Maurice Sililo Manyando, Dima Nazzal, Emily Awino Ogutu, Zoe Sakas, Francisco Castillo-Zunino, William Kilembe, Robert A. Bednarczyk, Matthew C. Freeman, the Vaccine Exemplars Research Consortium

**Affiliations:** Rollins School of Public Health, Emory University, Atlanta, GA, USA; Laney Graduate School, Emory University, Atlanta, GA, USA; Center for Family Health Research in Zambia, Lusaka, Zambia; College of Engineering, Georgia Institute of Technology, Atlanta, GA, USA; H. Milton Stewart School of Industrial and Systems Engineering, Georgia Institute of Technology, Atlanta, GA, USA; College of Arts and Sciences, Emory University, Atlanta, GA, USA; Emory University School of Medicine, Emory University, Atlanta, GA, USA; Biden School of Public Policy and Administration, University of Delaware, Newark, DE, USA; Yale School of Medicine, Yale University, New Haven, CT, USA; School of Public Policy, Georgia Institute of Technology, Atlanta, GA, USA

**Author notes:** Correspondence to; 1518 Clifton Road NE, Atlanta, GA, 30322. **Authors’ contributions:** KH, AE, WK, RB, MCF: project conceptualization and methodology; KM, CC, BD, CK, AL, MM, MSM, EAO, WK, RB: investigation and data curation; KM, KH, CC, RD, BD, AE, PK, AL, MM, MSM, DN, EAO, ZS, FCZ, WK, RB, MCF: formal analysis; KM, KH, RD, BD, AL, EAO, ZS, FCZ: writing - original; KM, KH, RD, AE, AL, ZS, FCZ, WK, RB, MCF: writing – review and editing; all authors provided approval of the submitted version. **Consortium author emails:**. **Consortium authors’ contributions:** NB, MD, CE, KI, BPM, WO, MR: project conceptualization; LB, SJ, CK, AM, HP, LP: investigation and data curation; LB, CE, KI, CJ, HJ, SJ, CK, RM, AM, WO, HP, LP, SR, VT, ZY: formal analysis; MD, CE, KI, RM, WO, SR, ZY: writing – review and editing; all authors provided approval of the submitted version.

**Keywords:** Vaccine policy, vaccine programming, childhood vaccination, health systems strengthening, implementation research

## Abstract

**Introduction:** The essential components of a vaccine delivery system are well-documented, but robust evidence on how and why the related processes and implementation strategies prove effective at driving coverage is not well-established. To address this gap, we identified critical success factors associated with advancing key policies and programs that may have led to the substantial changes in routine childhood immunization coverage in Zambia between 2000 and 2018.

**Methods:** We conducted mixed-methods research based on an evidence-based conceptual framework of core vaccine system requirements. Additional facilitators and barriers were explored at the national and subnational levels in Zambia. We conducted a thematic analysis grounded in implementation science frameworks to determine the critical success factors for improved vaccine coverage.

**Results:** The following success factors emerged: 1) the Inter-agency Coordinating Committee was strengthened for long-term engagement which, complemented by the Zambia Immunization Technical Advisory Group, is valued by the government and integrated into national-level decision-making; 2) the Ministry of Health improved the coordination of data collection and review for informed decision-making across all levels; 3) Regional multi-actor committees identified development priorities, strategies, and funding, and iteratively adjusted policies to account for facilitators, barriers, and lessons learned; 4) Vaccine messaging was disseminated through multiple channels, including the media and community leaders, increasing trust in the government by community members; 5) The Zambia Ministry of Health and Churches Health Association of Zambia formalized a long-term organizational relationship to leverage the strengths of faith-based organizations; and 6) Neighborhood Health Committees spearheaded community-driven strategies via community action planning and ultimately strengthened the link between communities and health facilities.

**Conclusion:** Broader health systems strengthening and strong partnerships between various levels of the government, communities, and external organizations were critical factors that accelerated vaccine coverage in Zambia. These partnerships were leveraged to strengthen the overall health system and healthcare governance.

**Highlights:**

- This paper describes how policies and programs contributed to improved vaccine coverage in Zambia
- Communication, coordination, and collaboration between implementing levels were imperative
- Adjacent successes in health systems strengthening and governance were leveraged
- Policies in Zambia include flexibility in implementation for tailored approaches in each district

## 1. Introduction

Vaccination, a pillar of preventive healthcare, is one of the most influential and comprehensive public health interventions of the last century, averting an estimated 2-3 million deaths annually [1, 2]. Effective vaccination programs and the goal to attain universal access to safe, effective, and affordable vaccines by 2030 underpin fourteen of the seventeen Sustainable Development Goals [3]. The Global Vaccine Action Plan (GVAP) targets at least 90% country-level coverage of the third dose of diphtheria, tetanus, pertussis vaccine (DTP3) among 1-year-old children, a globally recognized proxy for vaccination system performance, and 80% DTP3 coverage in countries’ subnational levels [4]. Global DTP3 coverage reached 83% in 2009, but has remained relatively stagnant since then. Only 65% of the World Health Organization (WHO) Member States achieved 90% or greater DTP3 immunization coverage in 2018 [5]. That same year, the African Region reported the lowest DTP3 coverage among the six WHO regions with 73% coverage, a slight improvement from 71% coverage in 2010 [6]. However, within the African Region, certain exemplar countries have outperformed their peers regarding high and sustainable immunization coverage providing an opportunity to examine the critical components of effective vaccine delivery.

The essential components of an effective vaccine delivery system are well established and include strong leadership and governance, healthcare financing, human resources, supply chain, and information systems [7]. The identified determinants of vaccine coverage include intent to vaccinate, community access, and health facility capacity [8]. However, evidence on how these components contribute to immunization program performance, the effect of multiple simultaneous initiatives, and actionable recommendations for implementation are lacking for specific country and regional contexts [7, 8]. The role of contextual factors such as health system capacity, national-level economic and political factors, and global health policies – essential characteristics for understanding the settings within which vaccine delivery systems operate – have seldom been reported [7]. Applying a positive deviance lens to the relative contributions of these essential health system components to increases in immunization coverage may identify transferable lessons and support actionable recommendations to improve national immunization coverage and increase vaccine equity [9].

While median DTP3 coverage in the African region increased from 52% to 73% between 2000 and 2018, nine countries in the region have achieved exemplary improvements in coverage as defined in a forthcoming Exemplars in Vaccine Delivery protocol paper (unpublished manuscript), reaching beyond 73%, and even 85%, in this same period [6]. Zambia further improved and maintained high DTP3 coverage, with 85% and 90% coverage in 2000 and 2018, respectively [6]. Zambia has maintained consistently high coverage of the first dose of DTP (DTP1) since the early 2000s, while simultaneously closing the gap between DTP1 and DTP3 from 2000 to 2017, with strong growth periods from 2007 to 2009 and 2013 to 2017 (Figure 1) [10].

**Figure 1:**
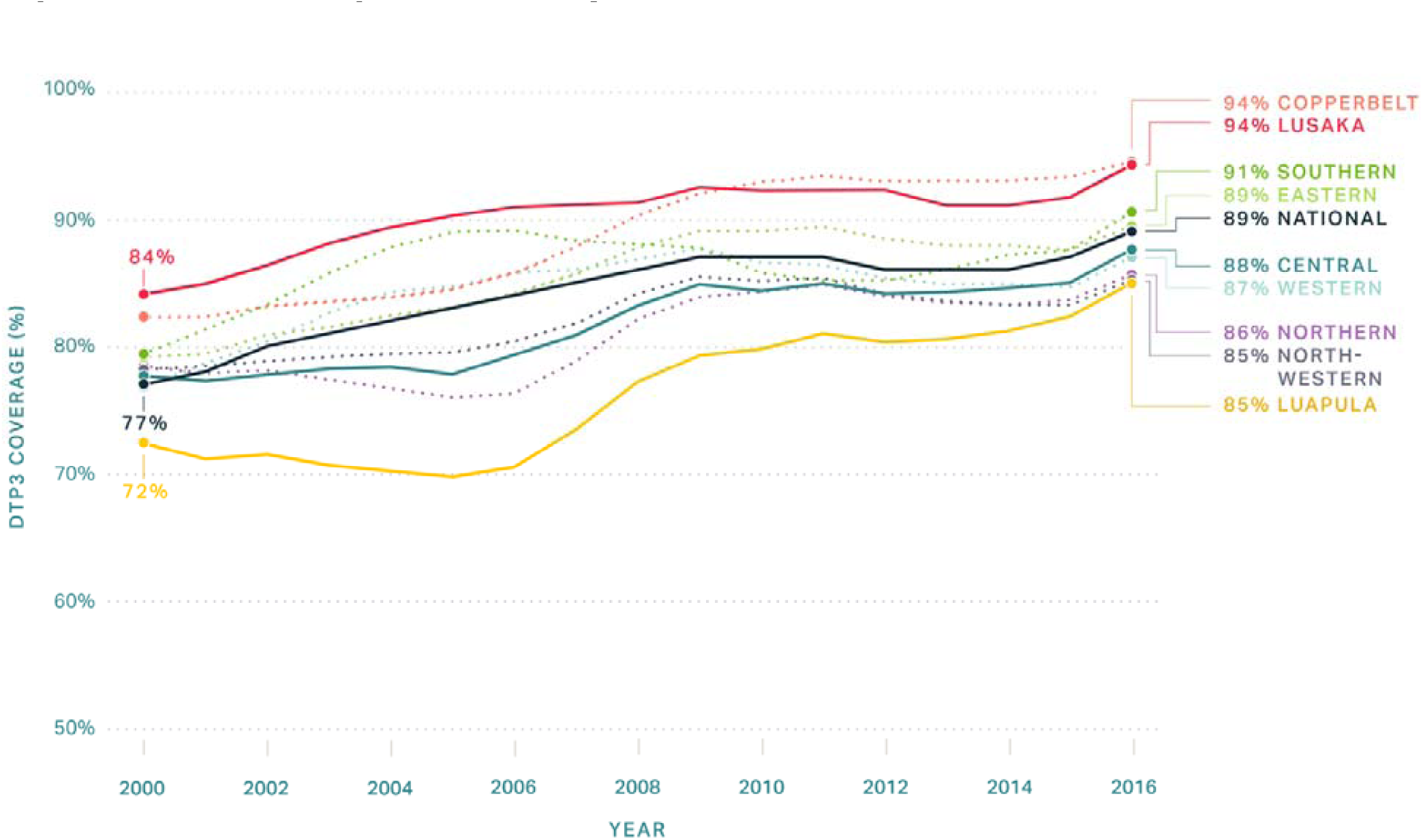
DTP3 coverage in Zambia by Province, 2000 – 2016 [1].

We used a positive deviance, case study methodology to identify and understand critical policies, programs, and other contributing factors that influenced high coverage rates in Zambia. Similar approaches have been used elsewhere to identify success factors associated with stunting, under-five mortality, and community health worker programs [9]. We modified an existing evidence-based conceptual framework that describes the core requirements for vaccine systems, and explored additional facilitators and barriers to vaccine coverage at national and subnational levels [8]. The framework incorporates demand and supply-side components to ascertain critical success factors and their implementation strategies. These results informed a novel framework that models the multidimensional critical components of an immunization program from public, private, and local entities that may support translatable lessons in other settings.

The goal of this study was to identify critical success factors that contributed to exemplary growth in routine early childhood vaccine coverage in Zambia that can lead to actionable recommendations for improving immunization programming in other settings. The critical success factors presented in this paper are not intended to be exhaustive or universal, nor to disregard other success factors or challenges; indeed, the context in which the program rests is crucial. Rather, this paper describes the implementation and execution of successful policies and programs and their means of contributing to the improvement in immunization program performance in the Zambian context.

## 2 Materials and methods

### 2.1. Study design and setting

We employed a hypothesis-generating, mixed-methods study design to explore the critical success factors that contributed to routine immunization coverage for children under 1 year of age in Zambia from 2000 to 2018. This case study was nested within the Exemplars in Vaccine Delivery Project of the Exemplars in Global Health Program, a project to identify components that made the study countries – Zambia, Nepal, and Senegal – exemplary in improving vaccine program performance [9]. Discussion of additional success factors, challenges, and contextual factors are described elsewhere [11].

We developed a conceptual model (Figure 2) to organize the complex interplay of factors impacting global childhood vaccine coverage, based on the work of Phillips et al. and LaFond et al., and a broader review of the vaccine confidence and coverage literature (Figure 2) [8, 12]. We conducted an initial scoping visit to gather preliminary information which guided research tool development. Conclusions about the role of specific practices and interventions were driven by empiric data collection and analysis rather than *a priori* assumptions about their importance.

**Figure 2.**
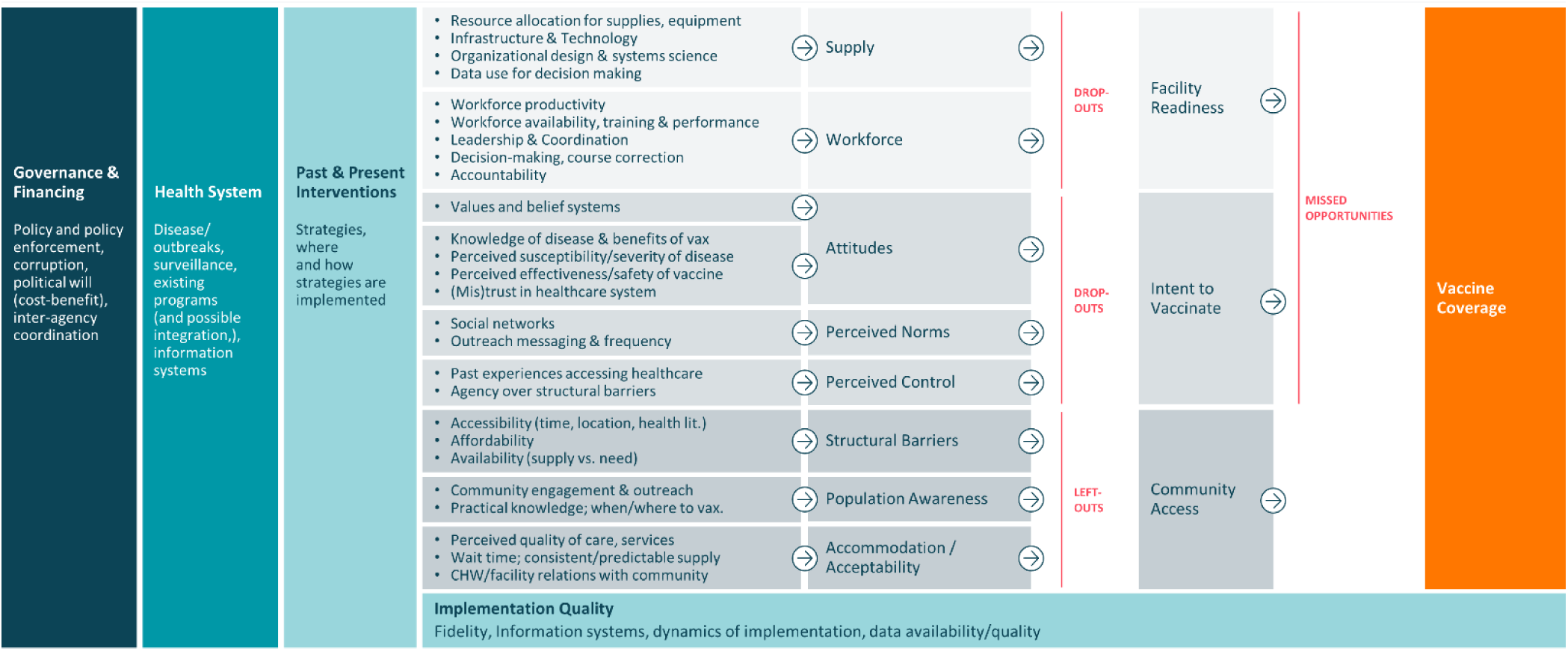
Conceptual framework of the drivers of vaccine delivery, derived from the scoping visit and Phillips et al [2].

Zambia was one of three countries selected as exemplars in vaccine delivery based on selection criteria as described in a forthcoming Exemplars in Vaccine Delivery protocol paper (unpublished manuscript). In the Zambian analysis, DTP1 and DTP3 coverage estimates served as proxies of the vaccine delivery system, with DTP1 as an index of access and DTP3 as an index of continuous utilization of healthcare services and the overall strength of the national immunization program [13]. In consultation with national stakeholders and available data, we selected three provinces in Zambia as study locations while considering heterogeneity via the following characteristics: 1) population density; 2) high and low performers, grouped by DTP3 coverage >90% and ≤90% coverage, respectively; 3) DTP3 coverage growth greater than 1% from 2000 to 2018; and 4) inclusion of the capital city of Lusaka.

Lusaka, Central, and Luapula Provinces were ultimately selected with rolling 3-year averages of 94.2%, 87.6%, and 85.0% DTP3 coverage in 2017, respectively [10]. We selected three districts within each province with consideration to varying DTP3 coverage and growth levels, the creation of new districts from 2011 to 2018, and stakeholder recommendations. Health facilities were selected based on the recommendations from the provincial and district health directors while considering differences in DTP3 coverage and growth. Additionally, a health facility was selected based on proximity to each district capital.

### 2.2 Quantitative data collection and analysis

Quantitative data were collected through secondary datasets, which included information from the Ministry of Health (MoH) and other partners. The primary goal of these analyses was to identify indicators that predict immunization coverage success among low- and lower-middle-income countries using cross-country and multi-year mixed-effects regression models to statistically test financial, development, demographic, and other country-level indicators. Financial results from this analysis demonstrate that government health spending was positively associated with improvements in immunization coverage [14].

### 2.3. Qualitative data collection and analysis

Qualitative data were collected between October 2019 and February 2020 at the national, provincial, district, health facility, and community levels. Data were collected by the Center for Family Health Research in Zambia (CFHRZ). The interview guides were informed by the Consolidated Framework for Implementation Research (CFIR) [15] and the Context and Implementation of Complex Interventions (CICI) framework [16]. Key informant interview (KII) and focus group discussion (FGD) guides were translated in Nyanja and Bemba languages by research assistants. All interview guides were piloted before use and adjusted iteratively throughout data collection. Research assistants conducted individual or group KIIs to identify key activities and strategies during change points seen in DTP1 and DTP3 coverage from 2000 to 2018. An initial list of KIIs was developed with CFHRZ and MoH officials then we used snowball sampling to identify additional key informants. FGDs were conducted to understand community-level factors immunization coverage and to identify the roles and responsibilities of frontline workers and community members within the immunization sector. Mothers, fathers, grandparents, and community-based volunteers (CBVs) were recruited from health facility catchment areas with the assistance of local health staff. The duration of KIIs and FGDs averaged one and a half hours. KIIs and FGDs were audio-recorded with the permission of participants. Research files, recordings, and transcriptions were de-identified and password protected. The activities are summarized in Table 1.

**Table 1:** Summary of research activities, October 2019 – February 2020.

| **Method** | **Participants** | **Number of activities** | **Number of participants** |
| --- | --- | --- | --- |
| Key Informant Interviews | Ministry of Health | 9 | 10 |
|  | Ministry of Education | 1 | 1 |
|  | Ministry of Finance | 1 | 1 |
|  | Partner organization | 11 | 15 |
|  | Provincial Health Office | 6 | 8 |
|  | District Health Offices | 10 | 19 |
|  | Nurses in-charge of health facilities | 7 | 10 |
|  | Community-based volunteers | 11 | 11 |
|  | Community leaders | 10 | 10 |
| Focus Group Discussions | Community-based volunteers | 10 | 60 |
|  | Mothers | 8 | 48 |
|  | Fathers | 1 | 6 |
|  | Grandparents | 3 | 18 |
| **Total** |  | **88** | **217** |

A theory-informed thematic analysis of the transcripts identified the contributing factors to success in the immunization program. We developed a codebook using a deductive approach and *a priori* data analysis, applying constructs from the CFIR and CICI frameworks, and adjusted the codebook based on the emerging themes. Transcripts were coded and analyzed using MaxQDA2020 software (Berlin, Germany). All transcripts were coded, relevant themes were identified, and visual tools were used to illustrate the findings. We considered setting and participant roles while identifying key points, and further contextualized data using historical documents and a literature review.

### 2.4. Ethical approval

The study was approved by the University of Zambia Biomedical Research Ethics Committee (Federal Assurance No. FWA00000338, REF. No. 166-2019), the National Health Research Authority in Zambia, and the Institutional Review Board committee of Emory University, Atlanta, Georgia, USA (IRB00111474). All participants provided informed consent, either written or with their thumbprint in the presence of a witness after the possible consequences of this study was fully explained.

## 3. Results and Discussion

KIIs were conducted at national (N=22) and subnational (N=45) levels; 21 FGDs were conducted with CBVs (N=10), mothers (N=8), fathers (N=1), and grandparents (N=3) within nine districts in Zambia. Table 2 provides the general demographic information of FGD participants.

**Table 2.** Demographic characteristics of focus group discussion participants. *Mean rounded to the nearest whole number

| **Characteristics** | **CBVs**<br>**(n=48)** | **Mothers**<br>**(n=36)** | **Fathers**<br>**(n=6)** | **Grandparents**<br>**(n=18)** | **Total**<br>**(n=108)** |
| --- | --- | --- | --- | --- | --- |
| Age* (range) | 46 (30-64) | 34 (18-51) | 47 (30-67) | 59 (51-69) | 47 (18-69) |
| Number of children* | 4 | 4 | 5 | 5 | 5 |
| Number of children in the home* | 4 | 4 | 3 | 5 | 4 |
| Age of youngest child vaccine decisions are made for* | 3 | 2 | 3 | 4 | 3 |
| Highest level of education, n (%) |  |  |  |  |  |
| No formal schooling | 0 (0) | 0 (0) | 0 (0) | 0 (0) | 0 (0) |
| Completed primary school | 30 (63) | 25 (69) | 4 (67) | 17 (94) | 76 (70) |
| Completed secondary school | 15 (31) | 11 (31) | 2 (33) | 1 (6) | 29 (27) |
| Completed post-secondary education (Bachelor's or Master's) | 2 (4) | 0 (0) | 0 (0) | 0 (0) | 2 (2) |
| Religion, n (%) |  |  |  |  |  |
| Islam | 2 (4) | 2 (6) | 0 (0) | 0 (0) | 4 (4) |
| Catholicism | 9 (19) | 2 (6) | 0 (0) | 2 (11) | 13 (12) |
| Protestantism | 24 (50) | 18 (50) | 5 (83) | 8 (44) | 55 (51) |
| Other Christianity | 13 (27) | 14 (39) | 1 (17) | 6 (33) | 34 (32) |

Through KIIs and policy reviews, we identified several government policies, health campaigns, disease outbreaks, natural disasters, and key interventions that may have affected immunization coverage in Zambia from 2000 to 2018. Figure 3 shows the timeline summarizing these factors as compared to the coverage of BCG, DTP1, DTP3, MCV1, and Pol3 vaccines in Zambia from WHO UNICEF Estimates of National Immunization Coverage (WUENIC) and Institute for Health Metrics and Evaluation estimates [6, 10]. Funds from Gavi, the Vaccine Alliance (“Gavi”) are also included in this assessment, as local governments are required to improve vaccine logistics and management to receive assistance. Immunization coverage fluctuated considerably from 2000 to 2018 for the above-listed vaccines, especially from 2006 to 2012. Based on DTP3 coverage, we identified five different time periods:

- Slow decrease in coverage from 2000 to 2007

⍰ Swift increase from 2007 to 2009
⍰ Swift decrease from 2010 to 2012
⍰ Steady increase from 2013 to 2017
⍰ Small decrease in 2018

**Figure 3.**
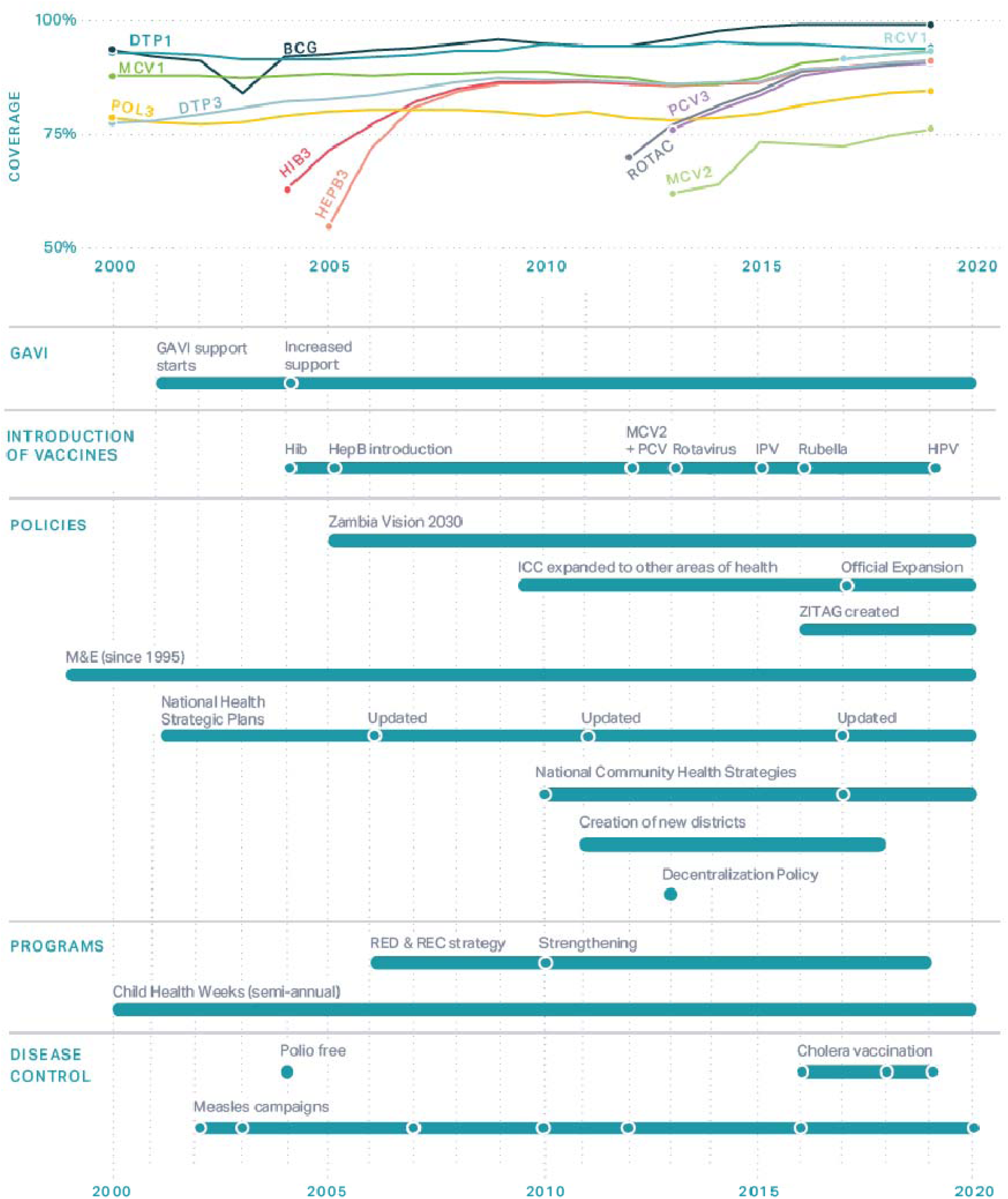
Immunization coverage with annotated events in Zambia, 2000 – 2020 [3, 4]. ICC = Inter-agency Coordinating Committee; ZITAG = Zambia Immunization Technical Advisory Group; M&E = monitoring and evaluation; RED = Reaching Every District; REC = Reaching Every Child.

Below, we describe specific strategies and policies that key informants described as success factors in Zambia.

Success factors represent either endogenous innovations – homegrown innovations developed specifically within Zambia – or exogenous innovations – adaptions of global policies and approaches to the context in Zambia. While context and culture are key factors in the successes and challenges Zambia faced, this analysis focuses on the deliberate policies and statutes that formalized and operationalized the contributing factors that led to robust immunization coverage.

Our analysis led to developing a conceptual framework (Figure 4) that categorizes critical factors, or mechanisms of success, that contributed to improvements in immunization coverage in Zambia – communication, coordination, and collaboration. Within the “Conceptual model of the drivers of vaccine coverage,” we explored the elements that were unmapped within the “Governance and Financing,” “Healthcare system,” and “Past and Present Interventions” components of our conceptual framework (Figure 2) specific to the context in Zambia. We determined that successful pathways of action (arrows) primarily travel along policy/statutory routes or cultural routes. Though few routes are exclusively policy/statutory or cultural, this differentiation highlights the importance of local context in understanding historical successes in early childhood immunization. This framework shows the flow of critical factors between and within the levels of implementation in Zambia. The functional definitions for the mechanisms of success and levels of implementation are found in Table 3. The purpose of this framework is not to build an immunization program from the ground-up, but rather to identify strategies to reach higher immunization coverage.

**Figure 4.**
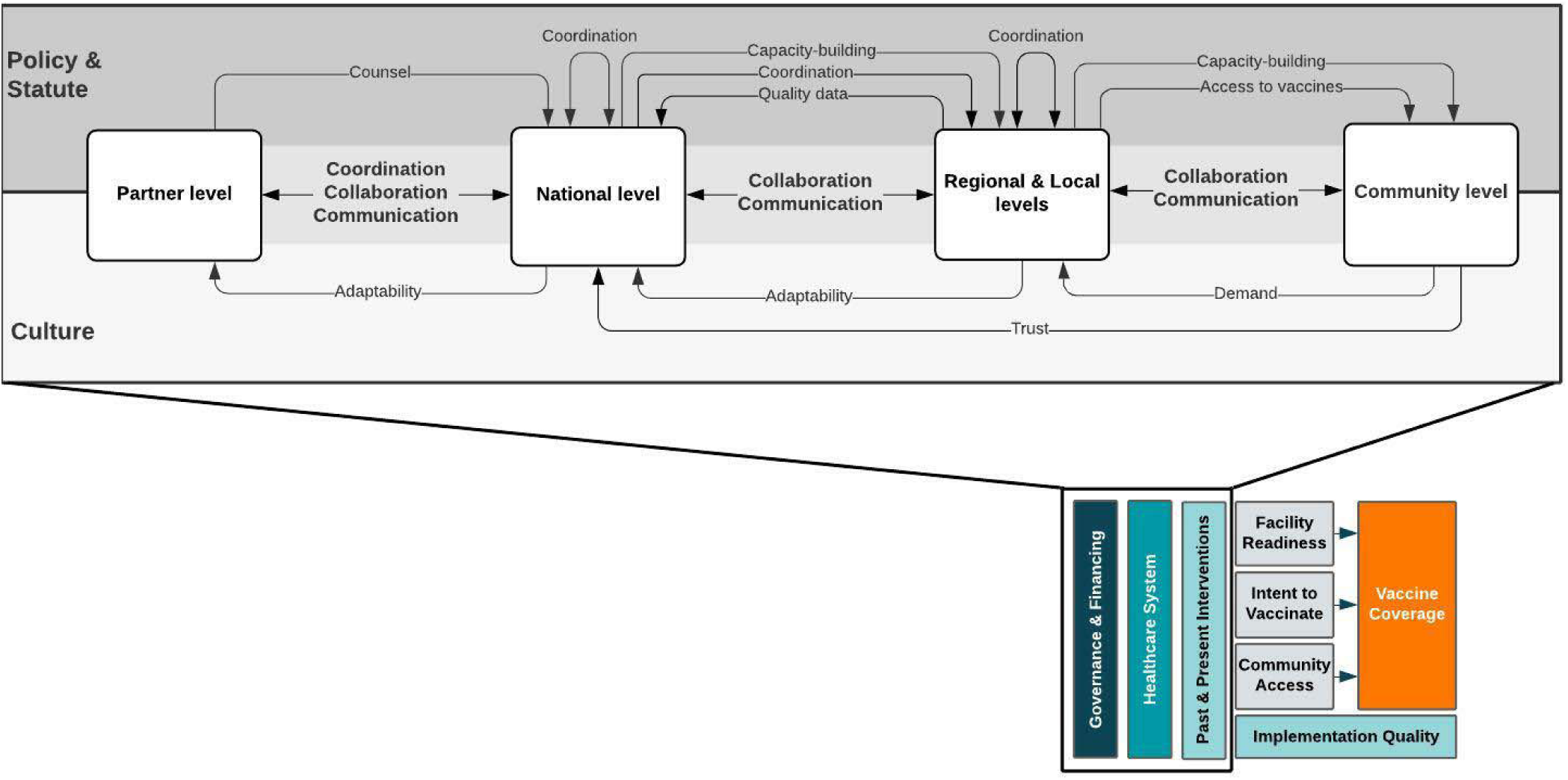
Critical factors that contributed to high coverage of routine vaccinations for children under 1 year of age in Zambia. All arrows represent successful pathways of action. Arrows flowing adjacent to “Policy & Statute” depict regulatory policies that Zambia operationalized into specific strategies. Arrows flowing adjacent to “Culture” depict more informal attributes that were associated with normative and historical context. Arrows flowing between the boxes illustrate the functional components across multiple levels of an exemplar country’s vaccine system that may have single directionality or bidirectionality.

**Table 3.** Functional definitions in the ‘Critical factors that contributed to high coverage of routine vaccinations among 1-year-olds in Zambia’ conceptual framework.

| **Mechanisms of success** | **Definition** |
| --- | --- |
| **Coordination** | The alignment of priorities between and within levels |
| **Communication** | Formal channels for input and feedback on priorities, data, and activities |
| **Collaboration** | Effective working relationships between levels towards the same priority |
| **Levels of implementation** | **Definition** |
| **Partner level** | Includes international and country-based organizations that support Zambia’s Expanded Program on Immunization |
| **National level** | Includes the ministerial offices that mandate the overall policy and provision of healthcare services |
| **Regional and Local levels** | Include provinces, districts, and health facilities that are responsible for the implementation and supervision of health promotion, curative, and preventive services at hospitals, health centers, and health posts |
| **Community level** | Population-based practices led by local stakeholders including community leaders (traditional, civic, and religious) and community members |

The Zambia Vision 2030, implemented in 2006, is a key policy that coheres long-term development agendas to the socioeconomic goals of the country and identifies equitable access to quality healthcare as a national priority. Informed by lessons learned from other countries, a series of strategic health plans stemmed from the Zambia Vision 2030 and seem to drive implementation decisions to improved coordination, collaboration, and communication across ministries, partners, and donors to promote stewardship over national policies [17].

### 3.1. Coordination mechanism

#### 3.1.1. Established distinct functions and relations between the ICC and NITAG for complementary long-term mandates

Inter-agency Coordinating Committees (ICCs) were initially created in the 1990s by WHO as a country-level working group to coordinate polio eradication programming [18]. In 2006, Gavi adopted the ICC as a forum to support the management of Gavi funds for immunization programming, requiring involvement from key MoH personnel and in-country partners [19]. International guidelines recommended that each country establish a National Immunization Technical Advisory Group (NITAG) as an independent advisory group to facilitate rigorous decision-making at the national level and present context-specific evidence to inform policy and program development [20]. The technical advisory role of the NITAG complements the ICC’s coordination of operational components of the immunization program and the strategic direction of other health sectors.

Zambia strengthened its ICC and NITAG for long-term engagement between the entities and within national-level planning for the immunization program. The Zambian ICC diversified internal stakeholders to include more technical experts, private partners, community organizations, external donors, and subnational representatives. Additionally, the ICC expanded its scope to other health sectors in addition to immunization, allowing for horizontal coordination and operation. For high functionality, a NITAG should have clearly defined and distinct roles from the ICC, and acceptance from the national government [21]. In Zambia, the ICC and NITAG have formal interactions with defined roles and responsibilities, in which the NITAG presents evidence supporting recommendations on a certain topic and the ICC considers the evidence for decision-making. Zambia decision-makers value the NITAG as a *“key component of the governance system”* (Ministry of Health Officer) that complements the ICC’s decision-making by providing context-specific recommendations [22]. The distinct roles between the ICC and NITAG accommodate improved integration of both bodies within national decision-making, a vital component for the success of a NITAG [20, 21]. This is a component of the coordination mechanism of success as the alignment of priorities towards a common strategic vision enhanced vaccine program performance. This collaborative approach will likely continue to support future improvements and longevity of immunization policies, service delivery, and consequently, vaccinated children.

#### 3.1.2. Employed a frontline approach to monitoring immunization coverage, leading to informed and timely decision-making at the subnational level

Frontline health workers in Zambia gained localized ownership of decision-making through the routine collection and review of community data. Health facility staff use this data to identify the areas with lower immunization coverage to establish outreach sites, discuss reasons for dropouts, calculate the coverage rates at both static and outreach immunization posts, determine indicators to monitor and evaluate outreach activities, estimate resources and funding needed for outreach activities, and assess progress towards health targets. Data include the following: 1) the number of outreach posts, 2) the number of mobile posts, 3) the number of CBVs, 4) headcounts of children 0-11 months and 1-5 years, 5) community member deaths, and 6) identification of leaders in hard-to-reach areas.

To improve and sustain the quality of data reporting and management, the Monitoring and Evaluation Department at the national level provides trainings to staff at all levels on health information systems and tools. Integrated meetings for data review are held at the district and provincial levels monthly and quarterly, respectively, to monitor health worker performance, review indicators, and make timely decisions to address gaps in immunization coverage. Community-level stakeholders may attend these meetings to discuss challenges, lessons learned, or provide recommendations for improvements. The MoH formalized protocols for data review and encouraged possession and responsibility of the data at all levels – with an emphasis on the health facility and community levels so that there is greater transparency and alignment of priorities within and across levels, a component of the coordination mechanism of success [23-28].

> *“We have been advising facilities that they need to own the data, they have to make sure that they utilize it by analyzing*… *If they are not meeting their targets, they need to assess themselves and see where they are going wrong… For those [health facilities] that are performing well, we find out what they are doing and we try to replicate to the other facilities that are not doing well*.*” District Health Officer*

In the last decade, Zambia emphasized the use of data for decision-making, especially at subnational levels, to assess equitable access to services and then implemented various interventions to improve data quality. District-level monitoring and evaluation of data have become increasingly common, as recommended by WHO, and routine monthly and quarterly reviews of health facility and district-level data have contributed to improved immunization program performance [7, 29-31]. Zambia went further than these recommendations by delegating specific roles at all levels – health information officers at the provincial and district health offices, environmental health technicians and community health assistants in health facilities, and neighborhood health committees (NHCs) in communities – as responsible for collecting, reviewing, and reporting data on a routine basis, which promoted evidenced-based decision-making in district and community action planning. Zambia also simplified data collection tools and indicators to promote the utilization for local decision-making, a practice also seen in Kenya and Mozambique [30, 32]. According to key informants, these interventions have been critical to timely data review to make iterative adjustments for immunization performance improvement.

### 3.2. Communication mechanism

#### 3.2.1. Operationalized bottom-up feedback to adapt policies and programs to the local context

GVAP called for comprehensive national immunization plans developed through a bottom-up process that includes key stakeholders from all national, subnational, and community levels [4]. Previous research has explored the importance of district health management teams tailoring immunization services to local contexts to improve coverage; however, few studies describe the steps for execution [7, 12, 31]. Global policies and guidelines are expected to be adapted to country context as necessary, and immunization programming is most successful as a shared responsibility among the national, subnational, and community levels [7]. Community ownership and trust in the government are built through strong collaboration and communication with the district health teams and local government, but it is unclear which strategies have been successful in various countries [12].

In Zambia, the national government instituted transparent policymaking in the Zambia Vision 2030, and formalized channels for feedback from the bottom up to ensure integrated planning and budgeting, a communication mechanism of success [17]. Through decentralization, while the MoH at the national level focuses on policymaking, implementation and management of immunization programming shifted to the subnational levels. The Zambian model to promote collaboration and communication between levels includes the District Development and Provincial Development Coordinating Committees. These regional multi-actor platforms identify development priorities and estimate funds required for implementing interventions. These platforms then submit their recommendations to the national level for review and approval. Additionally, the provincial level ensures that national policies are implemented and provides supportive supervision to the implementing districts and health facilities. As a strategic objective of GVAP, the provincial health offices undergo performance reviews for accountability to monitor and evaluate the implementation of policies [4]. When challenges arise at the subnational level, national policies may be adapted. District and provincial health offices host meetings for community representatives to participate in the planning and evaluation of frontline activities and provide suggestions for changes. Community stakeholder feedback is incorporated into future policy iterations to create adapted approaches to the local context. Ultimately, providing an opportunity for iterative adaptation of policies improved subnational and community stakeholder ownership of program planning, implementation, and concordance across levels.

> *“Once you have identified that there is actually an issue that needs attention, then these policies are revised and of course updated so that they match what is current. [Policies] are more or less like living documents to match the prevailing circumstances… Our planning has improved quite a lot because it’s now bottom-up. So, we are the ones that have quite a lot of input in these documents*.*” Provincial Health Officer*

#### 3.2.2. Created a well-structured communication system to engage the public in immunization messaging via multiple channels

The Zambia Vision 2030 policy laid the groundwork for a partnership with the media (e.g., television, radio stations), and described information as a “resource” critical for socioeconomic development [17]. At the national level, the Department of Health Promotions, Environmental Health, and Social Determinants within the MoH collaborates with media organizations to disseminate accurate and timely information. The MoH orients media personnel on the introduction of new vaccines and immunization campaigns and educates them on vaccine importance. At the subnational levels, provincial media liaison officers and district health promotion officers engage local radio stations across different sectors. Provincial and district health officers, and occasionally community leaders, are invited to local radio stations and are provided free airtime several times per year to discuss immunizations. This arrangement provides the media the influence to dispel myths and misconceptions related to vaccinations.

> *“When the program is starting, the media personnel would be called and then they are sensitized and given education about the program, like immunization*… *There were certain myths that were associated with immunization from the communities… The media has played a critical role to dispel some of those myths and make the communities understand, actually to say that vaccines are good for the children*.*” District Health Officer*

Open communication between community leaders and MoH officials at the district and regional levels has shaped community views on vaccines. The key development and health policies in Zambia reference training and engagement of community leaders as essential components of vaccine delivery [24-27]. Both community leaders and subnational ministry officials noted that vaccine delivery has always required the approval and participation of traditional leaders, who act as gatekeepers to their communities. The MoH holds indabas – formal community meetings – at least once per year at the provincial level with traditional and religious leaders. These meetings often provide education on community health, including vaccinations, address prominent vaccination misconceptions, and provide leaders the opportunity to discuss challenges in their communities.

Additionally, health facility staff meet with community leaders every 1-3 months to discuss upcoming activities such as Child Health Week, introduction of new vaccines, and notification of disease outbreaks. Leaders may share challenges faced in their community and suggest solutions, and may report hesitant individuals to health staff, who can counsel them further. The involvement and input from traditional leaders in local activities contribute to community ownership, buy-in, and consistent vaccine service delivery, a communication mechanism of success [26, 27].

> *“We make sure that the community is fully engaged about the benefit of ensuring that their children are vaccinated, and we need to ensure that within the community, the gatekeepers are helping us*.*” Provincial Health Officer*

### 3.3. Collaboration mechanism

#### 3.3.1. Engaged a faith-based organization for healthcare service delivery and outreach

The Churches Health Association of Zambia (CHAZ) is a Christian faith coalition formed in 1970 to organize church health institutions and faith-led community-based organizations for a greater impact on health service delivery. In 1973, CHAZ signed a memorandum of understanding with the MoH, in which CHAZ would provide support for mission health institutions by leveraging the strengths of faith-based communities [33]. This long-term organizational relationship led CHAZ, in partnership with local faith-based and community-based organizations, to operate numerous hospitals, rural health centers, and community-based organizations throughout Zambia. Through Gavi Health Systems Strengthening support and other grants, CHAZ member institutions provide 40% of national and over 50% of rural healthcare services while the national-level government provides essential supplies to these facilities. These services are offered to anyone, regardless of religious belief. To maintain data transparency with the government and partners, CHAZ-run facilities report data through the standard Health Management Information System. CHAZ operations have been integrated into the public health system for greater accessibility to services for remote areas in particular. CHAZ and the MoH demonstrate an effective working relationship towards the same priority to increase access to healthcare, which exemplifies the collaboration mechanism of success.

CHAZ engages multiple platforms for message development and delivery. CHAZ is involved at the national level in immunization message development for the general population and the media, especially during the introduction of new vaccines and Child Health Week. CHAZ develops and presents concept notes for message development and delivery to the immunization technical working group to review and revise. CHAZ also has a champion program in which it identifies and trains media and community members to disseminate immunization messaging to reach minority religious groups. Finally, CHAZ works with community organizations and health facilities to ensure equitable health outreach and service provision across Zambia.

#### 3.3.2. Elevated the partnership between the community and health system through Neighborhood Health Committees (NHCs)

Community participation in outreach planning and implementation contributed to improved immunization coverage in other settings [7, 12]. Across all interviews, participants discussed community engagement and community-driven strategies to combat health challenges. CBVs in Zambia are a fundamental component of community health and their roles are integrated into the delivery of primary healthcare services. The Zambia National Health Strategic Plans and the National Community Health Worker strategies (2010 and 2019) formalized the responsibilities of frontline workers which includes their role in planning for community health activities [23-25, 27]. Formalizing the CBVs’ role in community-level decision-making promotes an effective relationship with the health facilities, a key component of the collaboration mechanism of success.

NHCs, a group of CBVs in communities in Zambia, institutionalized the link between communities and the health system. NHCs exist in 84% of communities, with more engagement in rural areas, and consist of community-appointed volunteers whose primary role is to coordinate and supervise other CBVs on behalf of the health facility staff [27]. NHCs are assigned specific responsibilities in the Reaching Every Child Strategy and draft microplans specific to their catchment zone, which are then combined with other health facility catchment zones to develop the community action plan. NHCs also conduct situation analyses to map hard-to-reach areas and explore reasons for non-vaccination [34]. Additionally, NHCs work with health facility staff to develop the outreach timetable, communication plan, and local leadership engagement plan. In Zambia, community members respect NHCs due to their appointment by community leaders and their link to the health facility in planning, evaluation, communication, and coordination.

> *“There is the NHC that spearheads all community activities because all the CBVs, all community volunteers, they report through the NHC*.*” District Health Officer*

### 3.4 Implications for investments in immunization programming

We found that many improvements in immunization coverage emerged from adjacent successes in health systems strengthening. Zambia adapted external immunization strategies to its context and leveraged changes in the health system and governance to accelerate coverage. Though some of these strategies may be unique to the context in Zambia (e.g., government structure), our findings regarding the success factors may have salience in other settings and apply to other health systems for a horizontal approach to healthcare.

Health systems strengthening has become a priority in recent decades as disruptions in health services due to natural disasters, disease outbreaks, or civil unrest expose weaknesses in a country’s health system [35]. Additionally, disease-specific public health programming may cause country governments to reallocate resources to a specific cause while drawing resources from other priorities [36]. The Immunization Agenda 2030 includes a strong focus on health systems strengthening, building on GVAP, and states that sustainable immunization programming should be embedded within primary health care [1]. WHO developed the Health System Building Block framework for health systems strengthening to enable improvements across multiple health sectors [37]; however, few studies describe practical approaches to implementing the building blocks of this framework [38]. Key components of the broader health systems strengthening were apparent when improvements in routine immunization program performance were described.

Additionally, we did not identify substantial differences in immunization programming between the three study provinces, which points to a streamlined program with strong governance. Differences in subnational coverage may lie in the contextual factors related to economic and socio-cultural considerations. Zambia’s policies and statutes include some flexibility in implementation to allow for tailored approaches in each district which influenced the communication and collaboration between levels. Lessons learned from horizontal approaches to maternal and child health and primary health care may be applied to the broader health system functionality, and thus immunization.

The experience in Zambia suggests that targeted vaccine interventions may have a short-term impact, but the sustainability of the immunization program is supported by consistent and reliable coordination, collaboration, and communication between the government, community, and partner networks as strengthened through health system reform. Deriving the key elements of these networks and how each contributed to immunization program performance is critical to determining how and why vaccine coverage improved in a specific context. Further research into the temporality of these networks and mechanisms of success as well as a prospective exploration of how changes in governance and health systems strengthening may impact immunization coverage would be beneficial.

This study has several limitations. First, we focused on Zambia as a positive deviant in vaccine exemplars but were unable to carry out a similar analysis in a non-exemplary country to compare immunization coverage. Second, the research tools focused on the factors that drove catalytic change and did not probe on interventions or policies that were unsuccessful. Third, using qualitative methods to understand historical events was challenging; interviewees often spoke about current experiences rather than discussing historical factors. However, research assistants probed respondents to reflect on longitudinal changes in the immunization program.

## 4. Conclusion

Our research revealed success factors related to coordination, communication, and collaboration led to improved immunization coverage and exemplary vaccine delivery in Zambia. Using a positive-deviant lens and mixed-methods research, our analysis allowed for a nuanced understanding of the various contributing factors to high and sustained coverage. Key informants in this study identified success factors related to strong partnerships between the government, communities, and external organizations for informed decision-making and context-specific policies and interventions. These findings are consistent with international recommendations and guidelines to bolster country immunization programs, while also highlighting specific steps Zambia took to adapt and implement immunization strategies to its context. The data indicate that Zambia leveraged key components from health system strengthening and governance to accelerate vaccine coverage. These findings could be adapted to other country contexts to considerable advantage, although more research is needed to better understand the salience and transferability of these components within other contexts.

## Data Availability

available upon request

## Abbreviations

CBV: Community-based volunteer
CFHRZ: Center for Family Health Research in Zambia
CFIR: Consolidated Framework for Implementation Research
CHAZ: Churches Health Association of Zambia
CICI: Context and implementation of complex interventions
ICC: Inter-agency Coordinating Committee NHC Neighborhood Health Committee
ZITAG: Zambia Immunization Technical Advisory Group

## Acknowledgements

We thank the Center for Family Health Research in Zambia for their partnership in this study. In addition, we thank Sarah Chesemore, Anna Rapp, Tove Ryan, and Ethan Wong from the Bill and Melinda Gates Foundation; Kate Buellesbach, Nathaniel Gerthe, Gloria Ikilezi, Caitlyn Mason, David Phillips, and Oliver Rothschild from Gates Ventures; and the Vaccine Exemplars Research Advisory Group for their insights, specifically Agnes Binagwaho, Laura Craw, Carolina Danovaro, Anuradha Gupta, Heidi Larson, Penelope Masumbu, Kate O’Brien, Helen Rees, Lora Shimp, and Aaron Wallace. We gratefully acknowledge the participants who gave their time and insights to help us better understand Zambia’s vaccine delivery system.

## Data statement

The data are not publicly available as all data are confidential.

